# Evaluating the Cultural Validity of the Montreal Cognitive Assessment: A Comparison of Performance in Bengali and English tests in Bangladeshi adults with Parkinson’s disease in East London

**DOI:** 10.64898/2026.02.18.26346549

**Authors:** Anisa J. Shahid, Sheena Waters, Mandeep Singh, Alexandra Zirra, Tahrina Haque, Essa Bhadra, Ellen Camboe, Brook Huxford, David A. Gallagher, Thomas Boyle, Caroline Budu, Charles R. Marshall, Alastair J. Noyce, Kamalesh C. Dey

## Abstract

**Background:** The Montreal Cognitive Assessment (MoCA) is a recommended brief screening tool to detect cognitive impairment in people with Parkinson’s disease (PD).

**Objective:** To compare English and Bengali MoCA performance in Bangladeshi individuals with PD in East London.

**Methods:** This cross-sectional study involved participants completing both English and Bengali MoCA. Analyses included ANCOVA, paired and unpaired t-tests, and Bland-Altman methods in full and age-matched samples.

**Results:** Fifty PD participants and 22 healthy controls (HC) were included in the full analysis. Both groups scored higher on Bengali than English MoCA (mean difference ∼4 points, p<0.001). Age-matched analyses (n= 29 PD and 22 HC) detected PD-control differences with the Bengali but not English version (p=0.02). Bengali scores aligned more closely with multidisciplinary assessments, though mean scores remained below normative cut-offs.

**Conclusion:** Bengali MoCA improves detection of cognitive differences over English but still overestimates impairment, supporting the need for culturally adapted tools and population-specific cut-offs.

## Introduction

Non-motor symptoms in Parkinson’s disease (PD) include cognitive impairment affecting attention and memory^1–4^. Parkinson’s disease dementia (PDD) is characterized by a progressive decline in cognitive function which significantly interferes with daily life^5^. Both PDD and mild cognitive impairment (MCI) are associated with increased disability, reduced independence, and greater caregiver burden, impacting quality of life ^4–6^.

In the Bangladeshi population, PD prevalence may be increasing^7,8^. Underdiagnosis or delayed diagnosis may occur in Bangladeshis due to limited awareness of neurological conditions and stigma, especially for cognitive impairment^9–13^. In East London, 33% of Tower Hamlets’ population identify as Bangladeshi, many being Bengali speakers^14^. The East London Parkinson’s Disease (ELPD) project examines PD in diverse populations, including Bengali/Bangladeshi participants^15^.

The Montreal Cognitive Assessment (MoCA) is a recommended tool for screening for cognitive impairment in patients with PD. Although validated in some South Asian populations, the MoCA’s generalizability to UK Bengalis remains unclear^15–18^. Even when language is addressed in the MoCA, cognitive assessments may not be culturally fair due to the lack of culturally adjusted cut-offs and differences in familiarity with pen and paper tasks^16–18^. Limited awareness of PD and its cognitive impact in minorities like the Bengali community may delay detection of MCI and PDD^8–11^. This study aimed to assess the cultural validity of the MoCA and agreement between the Bengali and English version of the MoCA in Bengali-speakers.

## Methods

### Study design, participants, and data collection

The East London Parkinson’s Disease (ELPD) project received ethical approval (18/SW/0255 and IRAS ID 242395), and all participants provided written consent. Within the ELPD project, a cross-sectional study assessing the MoCA’s cultural validity in Bangladeshi people with PD and healthy controls (HCs) was conducted.

All participants with PD were recruited from the Movement Disorders clinic at the Royal London Hospital (RLH), while HCs were recruited through clinics and community approaches in East London. The PD group included individuals self-identifying as Bangladeshi, with a clinical PD diagnosis according to MDS 2015 criteria^19^. The HCs comprised of individuals self-identifying as Bangladeshi, without PD or any neurological disease, without diagnosed cognitive impairment, and able to consent.

The MoCA tests were administered in the RLH or a home visit using both the English MoCA (version 8.3) and Bengali MoCA (version 7.1) conducted in an alternating order with 50% of each group receiving the English MoCA first. Both versions were completed in the same session with a short interval to minimize order effects. A one-point adjustment was applied for participants with fewer than 12 years of formal education, per MoCA scoring guidelines^20^. The Bengali MoCA included culturally appropriate naming and recall items. Demographic details were also recorded. All data were securely stored on Queen Mary University of London servers.

Cognitive status classification was determined using a gold standard multidisciplinary team (MDT) consensus diagnosis. The MDT, including cognitive and movement disorders neurologists, reviewed participant clinical data to establish a consensus classification of normal cognition, mild cognitive impairment (MCI), or Parkinson’s disease dementia (PDD). English and Bengali MoCA scores were compared to MDT clinical diagnostic categories, with a score of >25 indicating normal cognition, 19-25 indicating MCI, and <19 being classified as PDD^20,21^.

### Data analysis

Statistical analyses were completed with Python. This included a repeated measures analysis of covariance (RM ANCOVA), Welch’s t-tests, and Bland Altman plots. Samples were compared to assess between-group and within-group differences in MoCA performance across HC and PD groups in Bengali and English. Statistical significance was defined as p≤0.05. All data are presented as mean (95% confidence interval [CI]) unless stated otherwise.

To address the primary research question-whether test performance differs between the MoCA language versions, within-group comparisons were performed using a RM ANCOVA and included age as a covariate for the main analysis. Age and education level descriptive statistics were provided and compared between PD group and HCs using unpaired t-tests to analyze any significant differences. Age-matched analyses used ‘nearest-neighbor’ in Python matching without replacement, pairing 22 controls with 29 PD participants. For the within-group comparisons in the age matched cohort, a paired t-test was used when data were normally distributed and for between-group comparisons, Welch’s (unequal variances) t-tests were used as complementary analyses.

A Bland-Altman analysis was also conducted to assess agreement between English and Bengali MoCA scores within both age-matched groups (N=22 HC, 29 PD). This method assesses agreement between the two language versions and identifies any systematic bias.

## Results

Between May 2023 and December 2024, 72 participants (50 PD and 22 HC) were recruited (Supplementary Figure S1). The HCs were on average 10 years younger than the PD group (***p<0.01***), thus the RM ANCOVA included age as a covariate. An age-matched case-control subset (29 PD and 22 HC) was generated for comparison. Descriptive characteristics are reported (Table 1).

**Table 1.** Participants characteristics and MoCA score comparison between English and Bengali MoCA version in people with PD and HC (age-matched cohort).

| Variables | PD group (n=29) | HC group (n=22) | P Value |
| --- | --- | --- | --- |
| Age at assessment (year)<br>(mean±SD) | 57.8±9.8 | 55.0±10.5 | -- |
| Male (N, %) | 21(72.4) | 15 (68.2) | -- |
| Years of education (mean±SD) | 11.9±7.1 | 11.5±4.3 | -- |
| Age leaving school (mean±SD) | 18.7±9.0 | 18.4±5.2 | -- |
| English MoCA (mean, 95% CI) | 18.4 (16.3, 21.7) | 21.8 (19.2,24.4) | 0.06 |
| Bengali MoCA (mean, 95% CI) | 22.2 (19.7,25.1) | 25.9(24.2,27.6) | **0.02** |

Data presented as mean ± standard deviation, or N (%). MoCA data presented as mean (95% confidence interval); Statistically significant (p≤0.05) differences highlighted as bold.

In the full cohort, HCs scored higher than the PD group on the English MoCA (HC: M = 21.8; PD: M = 14.0), with a mean difference of −7.77 points (t (54.1) = −4.59, ***p<0.001***, 95% CI [−11.16, −4.38]). For the Bengali MoCA, the HCs again scored higher than the PD group (HC: M = 25.9; PD: M = 18.0), with a mean difference of −7.95 points (t (69.6) = −5.57, ***p<0.001***, 95% CI [−10.80, −5.10]) (Table S4). In the age-adjusted RM ANCOVA, group differences remained significant for the English (***p=0.03***) and Bengali (***p=0.02***) MoCA, and age was a strong negative predictor of performance (***p<0.001***).

In the age-matched analysis (29 PD, 22 HC), using the Bengali MoCA, the HCs scored significantly higher than the PD group (HC: M = 25.9; PD: M = 22.2), with a mean difference of (HC − PD) 3.70 points (t (49) = −2.36, ***p=0.02***, 95% CI [−6.85, −0.56]). Similarly, with the English MoCA HCs scored higher (HC: M = 21.8; PD: M = 18.4), although this difference did not reach statistical significance, with a mean difference of 3.4 points (t (49) = −1.90, ***p=0.06***, 95% CI [−6.92, 0.20]) (Table 1).

Within-group comparisons in this cohort revealed that Bengali MoCA scores were significantly higher than the English scores. In the PD group, scores were 3.79 points higher on average (t (28) = 7.24, ***p<0.001***, 95% CI [2.72, 4.87]). In the HC group, the Bengali MoCA scores exceeded the English scores by 4.14 points (t (21) = 4.83, ***p<0.001***, 95% CI [2.36, 5.92]).

Bland-Altman analysis showed a mean 4-point higher score on the Bengali MoCA, indicating systematic bias across scores (Figure S3).

### Cognitive impairment in PD group

In the full PD cohort (N=50), 43 participants had available MDT data. The MDT classified (N=43) 27.9% of participants as having normal cognition, 34.9% as MCI, and 37.2% as PDD. Using Bengali MoCA, 18.6% were classified as normal cognition, 32.6% as MCI, and 48.8% as PDD, whilst with the English MoCA there were lower percentages classified as normal and MCI, whilst a higher percentage was classified as PDD (Figure 1). Adjusted English MoCA scores (+4) improved the alignment of cognitive classification with the MDT, with a higher proportion of PD participants categorized as having normal cognition (16.3%) and fewer classified with PDD (44.2%) (Figure 1). Cognitive classification based on MoCA scores was also reported in the age-matched group (Figure S2).

**Figure 1.**
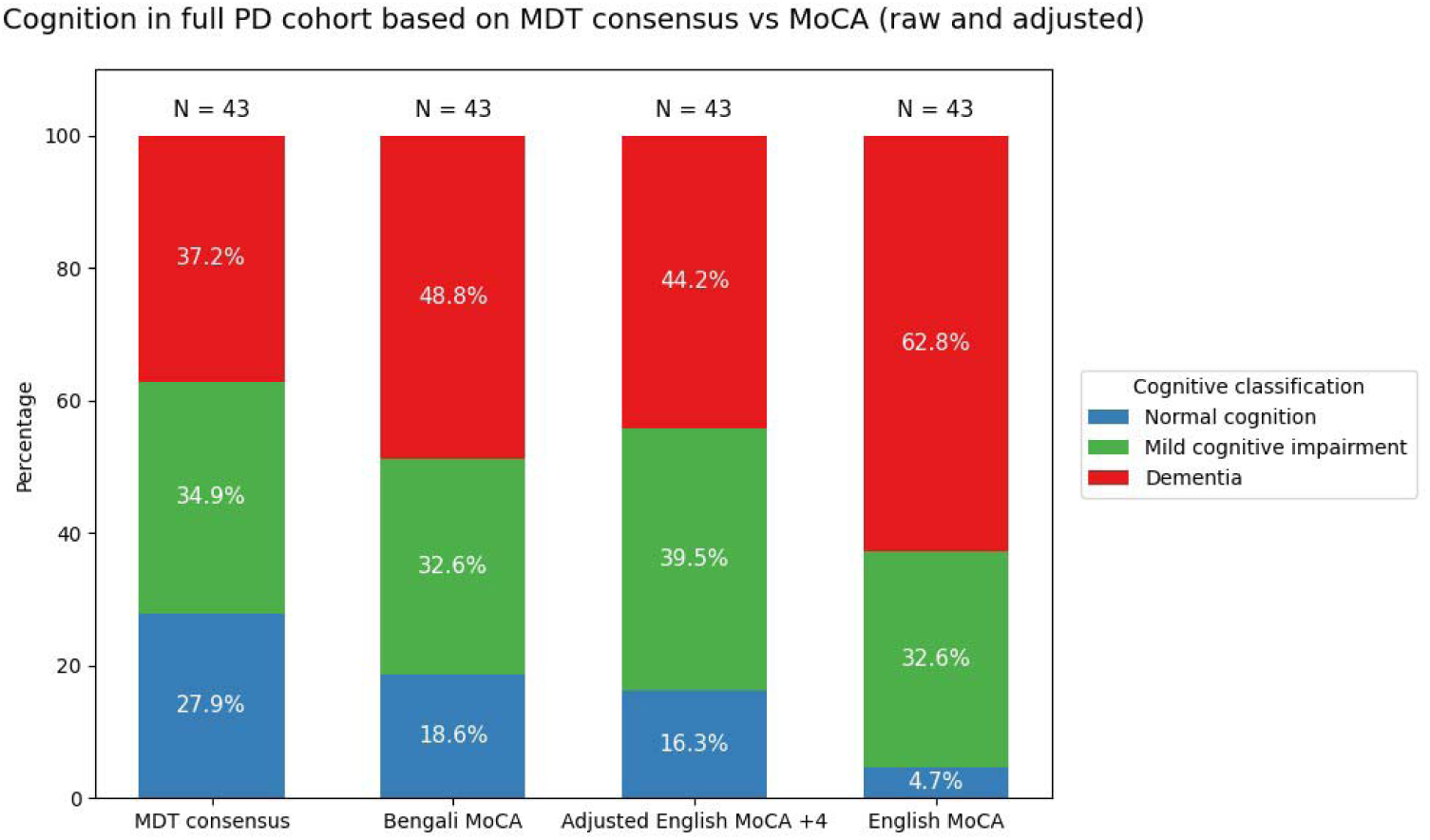
**Cognitive classification in people with Parkinson’s disease based on the MDT English MoCA scores and Bengali MoCA scores** (the stacked bar chart shows cognitive classification in the full PD cohort with available multidisciplinary team (MDT) consensus data (N = 43). The four bars represent cognitive status determined by: MDT consensus diagnosis, Bengali MoCA score thresholds, adjusted English MoCA thresholds (+4 points), and English MoCA score thresholds. Cognitive classifications are categorized as normal cognition (NC, blue), mild cognitive impairment (MCI, green), and Parkinson’s disease dementia (PDD, red). MoCA scores were classified as: <19 indicating PDD, 19-25 as MCI, and 25+ as NC. Compared to the MDT consensus, the English MoCA classified a greater proportion of participants as having PDD and fewer as cognitively normal, whereas the Bengali MoCA resulted in a higher proportion of participants classified as cognitively normal and fewer classified as having PDD but overall aligned more similarly to the MDT classifications. When English MoCA scores were adjusted positively by 4 points, an increase in alignment to the MDT and Bengali classification percentages is observed).

## Discussion

The culturally adapted Bengali MoCA was associated with higher scores across PD and control groups. As hypothesised, the HC group significantly outperformed the PD group on the Bengali MoCA, whilst this difference was less evident when using the English version. The lower mean MoCA scores in the full PD cohort, compared to the age-matched cohort, possibly reflect the inclusion of participants with more advanced cognitive impairment (PDD), some likely excluded during matching of the groups. The mean HC score on the Bengali MoCA (25.9) fell slightly below the standard cut-off of 26 for normal cognition^20^. Although the Bengali MoCA reduces linguistic and cultural bias relative to the English version, it may still overestimate cognitive impairment in this population.

Bland-Altman analysis showed that PD participants scored 3-4 points higher on the Bengali version, indicating it may be more appropriate for this population. These findings support that the English MoCA may require score adjustments in Bengali-speaking patients as it may underestimate cognitive ability due to linguistic and cultural biases.

This adjustment is further supported by categorical analyses comparing MoCA-based classifications with MDT consensus. The Bengali MoCA demonstrated closer alignment with MDT classification by identifying more PD participants as cognitively normal compared to the English MoCA. The Bengali MoCA also showed more similar distribution across mild cognitive impairment and dementia categories.

Adjusting the English MoCA by 4 points aligned classification more closely with the MDT consensus, reducing the potential over-classification of dementia and increasing concordance with clinical judgement. A higher proportion of PD participants were classified as cognitively normal when assessed in Bengali. Reliance on English-only assessments may therefore lead to misclassification and potential overdiagnosis of cognitive impairment in non-native speakers.

Education was accounted for in this study by adding an additional point for participants with fewer than 12 years of formal education due to its influence on MoCA performance, particularly in low literacy populations^21–23^. Age and other demographic factors influence cognitive decline in PD, and studies have underscored the need to consider these factors to reduce bias in comparisons between PD and control groups^5,6^.

Although the MoCA is regarded as a valid screening tool for early cognitive impairment, its sensitivity to cultural and linguistic factors remains a limitation^22–26^. Prior research notes such assessments may inadvertently measure cultural familiarity rather than underlying cognitive function^25–27^. Our findings extend this literature by demonstrating that culturally adapted versions of the MoCA improve raw performance scores but also alter diagnostic outcomes when compared against multidisciplinary clinical consensus, although residual cultural bias appears to remain even in the adapted version. Enhancing cultural validity in cognitive assessments can have implications for diagnostic accuracy, treatment planning, and equitable access to care.

The need for culturally adapted tools is particularly urgent in South Asian communities, where neurological research remains limited^9–13,15^. Cultural differences in symptom expression, disease understanding, and health-seeking behaviour impact quality of life for patients and caregivers^7,12^. Culturally sensitive diagnostic instruments are needed to avoid conflating cognitive impairment with linguistic, cultural, or educational background^24–27^. As a pilot study, our findings provide preliminary evidence for the utility of the Bengali MoCA in Bangladeshi populations. However, the small sample limits generalisability and larger age-matched cohorts are needed to confirm these findings across broader demographic and clinical subgroups

Overall, Bengali speakers perform better on the Bengali MoCA, which more closely reflected MDT cognitive classifications than the standard English version, regardless of PD status. These findings support culturally adapted assessments, and adjusted cut-off scores may improve classification and earlier detection of cognitive impairment in diverse groups.

## Data Availability

All data produced in the present study are available upon request to the authors.

## Acknowledgements

We express our sincere gratitude to all participants who generously contributed their time to this study. We acknowledge the contributions of all co-authors to the development of this manuscript. This work was supported by funding from the Michael J. Fox Foundation, to whom we extend our sincere thanks.

## Author’s roles

KCD and AJN designed the study protocol.

KCD, AJS, and MS recruited and assessed participants clinically.

AJS and SW performed the analysis. AJS, AJN, and KCD wrote the manuscript. All authors provided valuable feedback and approved the final version of the manuscript before submission.

## Supplementary figures and tables

**Figure S1.**
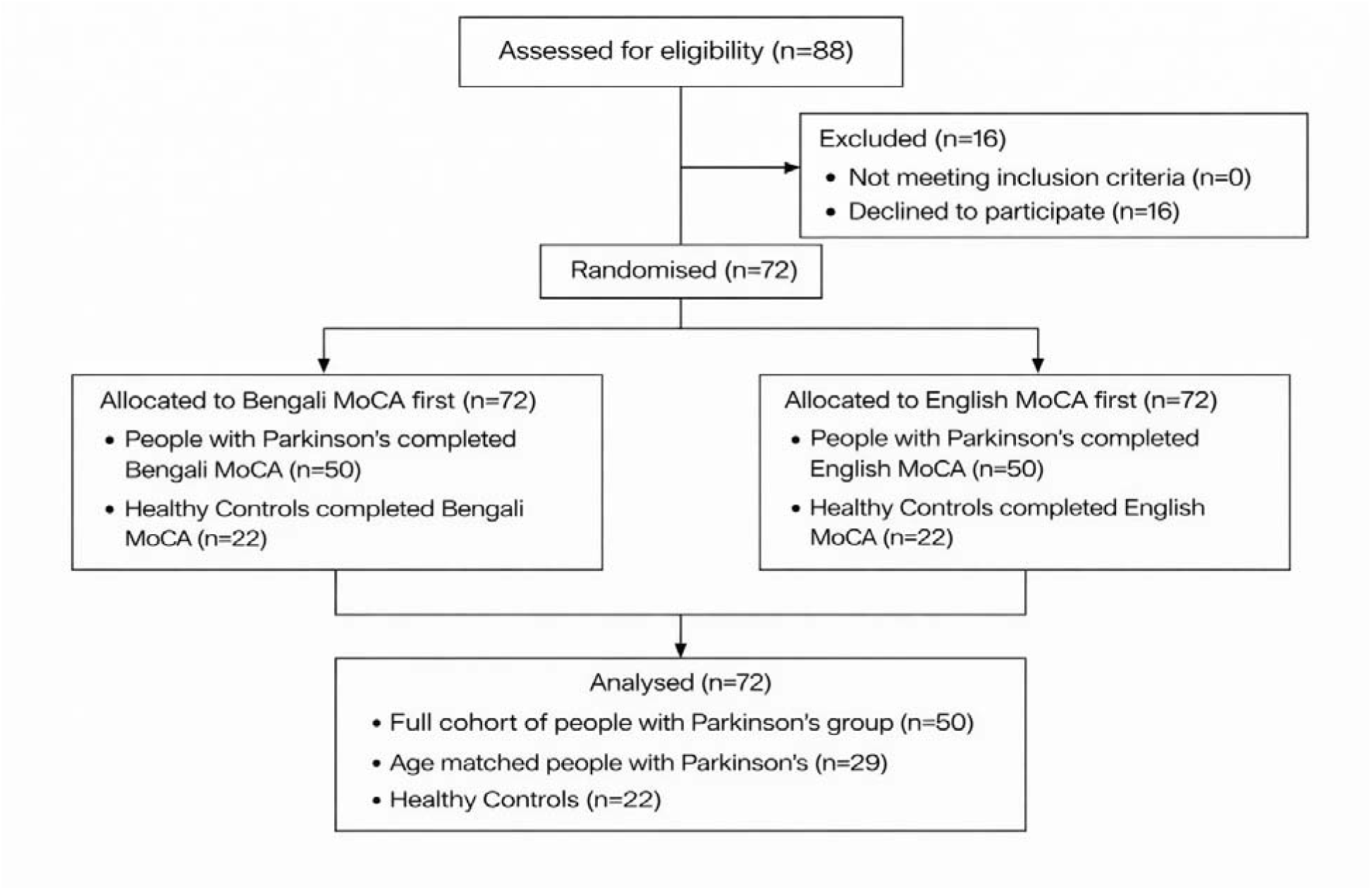
Participants flow throughout the study.

**Figure S2.**
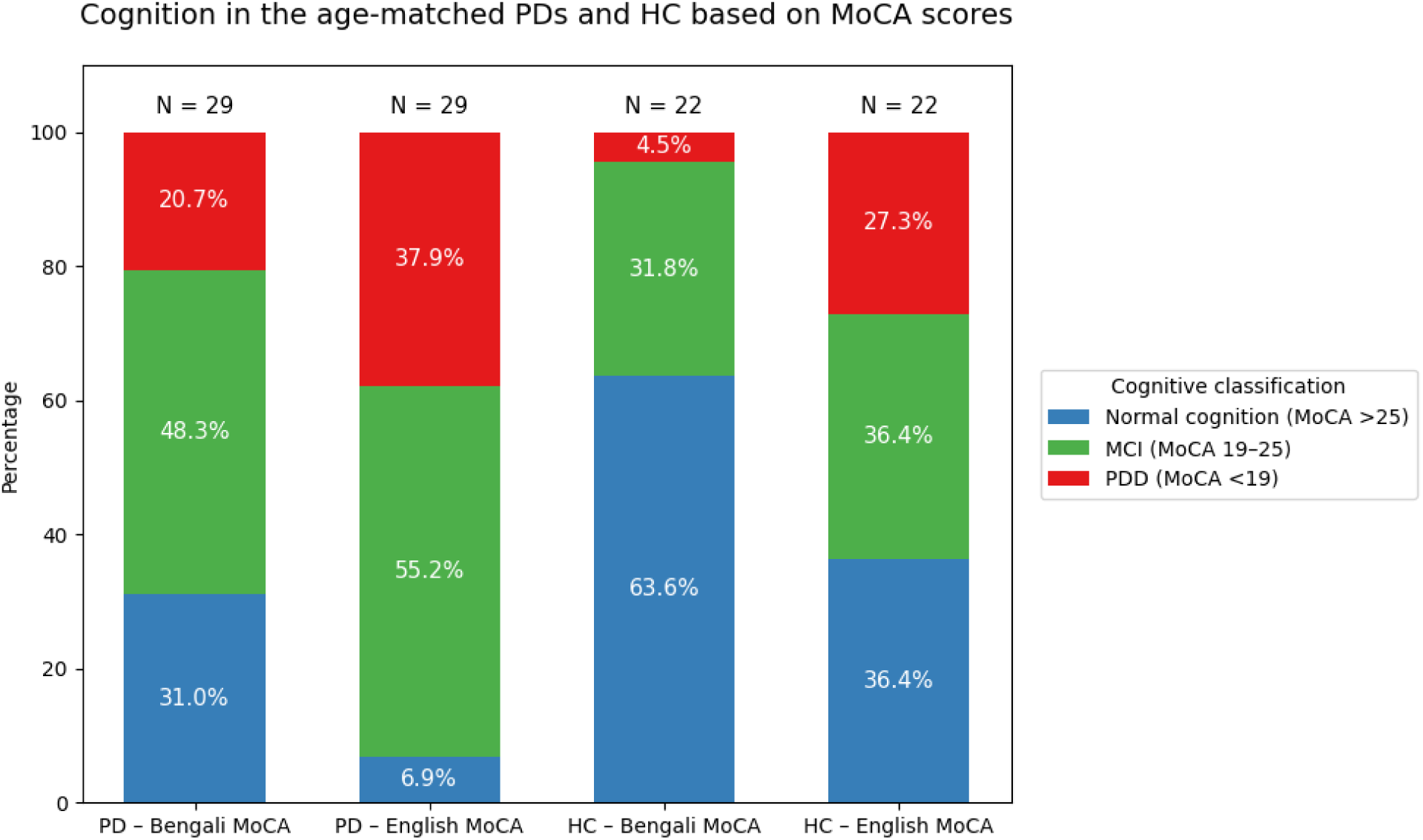
**Cognitive classification of the age-matched cohort (PD (N=29), HC(N=22) based on the English MoCA scores and Bengali MoCA scores** (the stacked bar chart shows cognitive classification in the age-matched PD and HC cohorts based on MoCA performance. The four bars represent cognitive status determined by Bengali MoCA scores in PD (N = 29), English MoCA scores in PD, Bengali MoCA scores in HC (N = 22), and English MoCA scores in HC. Cognitive classifications are categorized as normal cognition (NC, blue), mild cognitive impairment (MCI, green), and Parkinson’s disease dementia (PDD, red). MoCA scores were classified using standard thresholds: <19 indicating PDD, 19–25 as MCI, and ≥26 as NC.In the PD cohort, the Bengali MoCA classified a greater proportion of participants as cognitively normal and fewer as having PDD compared with the English MoCA, which identified a much higher proportion of participants in the dementia and MCI range. In the HC cohort, the Bengali MoCA yielded a higher proportion (63.6%) of individuals classified as cognitively normal and fewer classified as having PDD (4.5%) compared with the English version (NC=36.4%, PDD=27.3%). Across both PD and HC groups, the Bengali MoCA produced cognitive distributions consistent with higher cognitive performance relative to the English MoCA, supporting the influence of language and cultural factors on cognitive screening measures).

**Figure S3.**
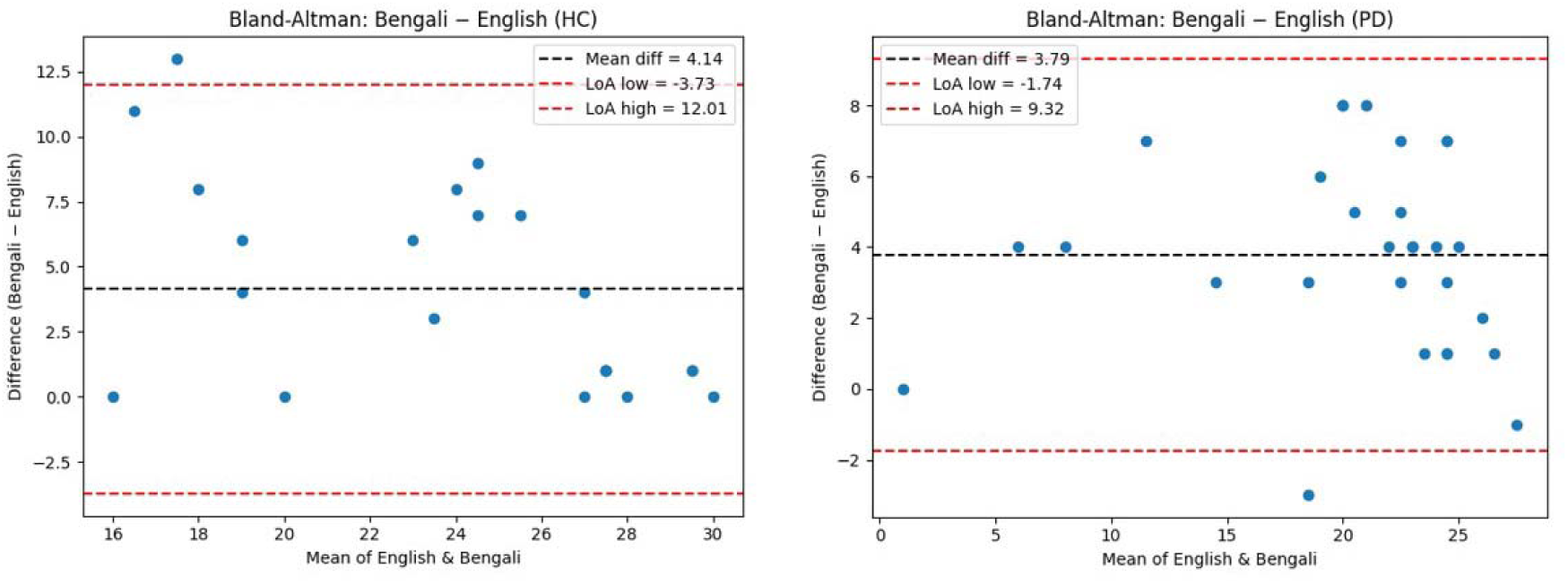
**Agreement between English and Bengali MoCA in HC and PD** (Bland–Altman plot illustrating agreement between total scores on the Bengali and English versions of the MoCA in participants with Parkinson’s disease (PD). The central dashed line indicates the mean difference between versions, demonstrating that PD participants scored on average 3–4 points higher on the Bengali MoCA compared with the English version. The upper and lower dashed lines represent the 95% limits of agreement, indicating variability in individual-level differences between the two versions across the range of scores. Bland–Altman plot illustrating agreement between Bengali and English MoCA total scores in healthy control (HC) participants. As in the PD group, HC participants scored consistently higher on the Bengali MoCA, with a mean difference of approximately 4 points. The limits of agreement demonstrate substantial variability, suggesting that while a systematic difference exists at a group level, individual scores differ sufficiently which limits direct interchangeability between MoCA versions).

**Table S4.**
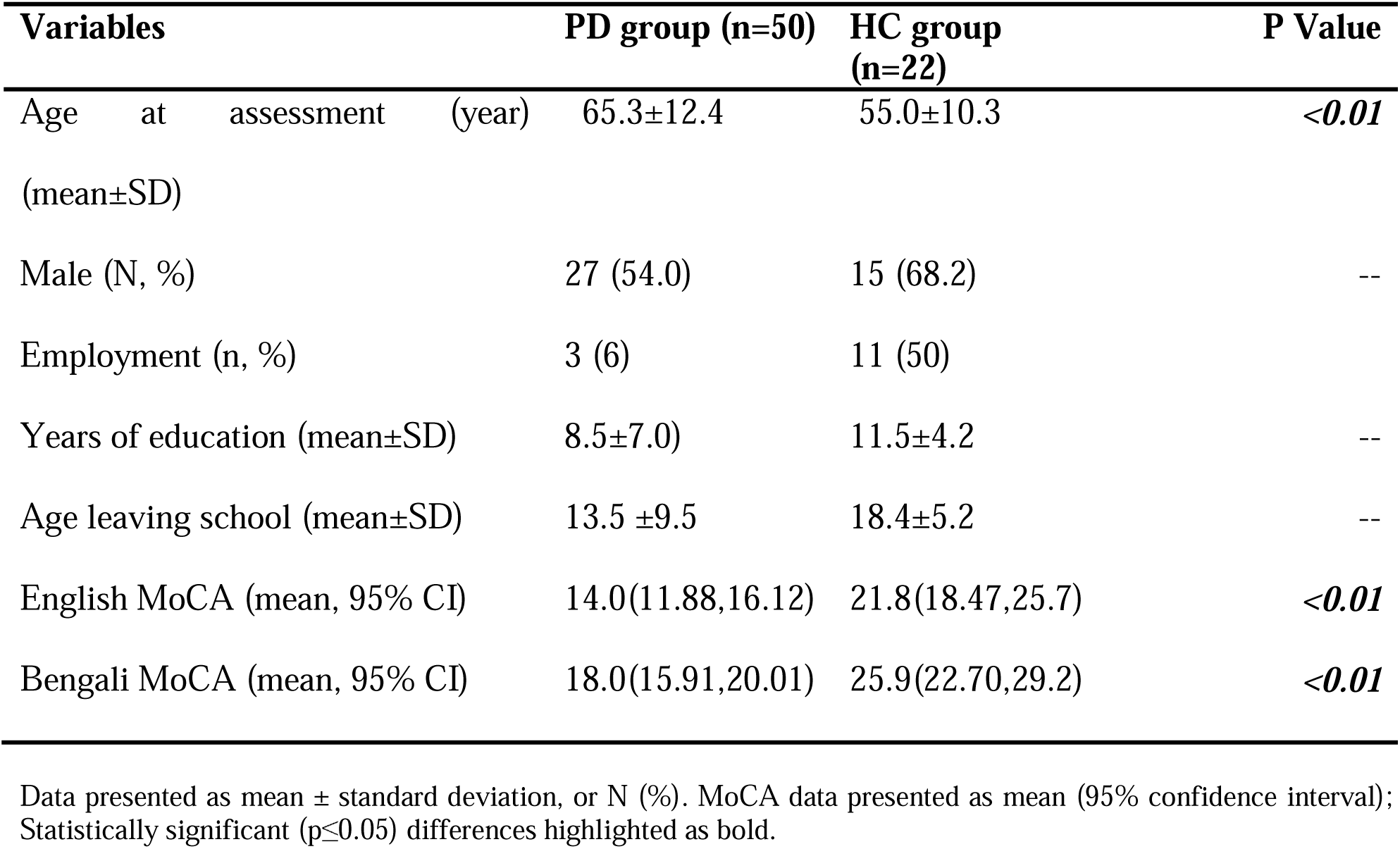
Full cohort characteristics.

